# In-clinic validation of a smartphone-based finger tapping test for use in neurodegenerative and neurological populations

**DOI:** 10.64898/2026.06.25.26356467

**Authors:** Morgan O’Connor, Mark E. Sanderson-Cimino, Zi Li, Sreya Dhanam, Anjali Sadarangani, Joshua Downer, Ray Fregly, Jack Taylor, Amy B. Wise, Kaitlin B. Casaletto, Leah K. Forsberg, Maria Luisa Gorno-Tempini, Hilary W. Heuer, Joel H. Kramer, John Kornak, Bruce Miller, Emily W. Paolillo, Riley Bove, Gil Rabinovici, William W. Seeley, Brad F Boeve, Howard J. Rosen, Adam L Boxer, Adam M. Staffaroni

**Affiliations:** Department of Epidemiology and Biostatistics, University of California, San Francisco, CA, USA; Department of Neurology, Weill Institute for Neurosciences, University of California, San Francisco, CA, USA; Department of Neurology, Mayo Clinic, Rochester, MN, USA

**Keywords:** frontotemporal dementia/frontotemporal lobar degeneration, progressive supranuclear palsy, motor speed and dexterity, digital biomarker, datacubed health

## Abstract

**Background:** Motor disturbances are common in neurologic and neurodegenerative syndromes. A standard motor speed and dexterity measure is the finger tapping test (FTT). The FTT has traditionally been administered in clinic using a mechanical FTT, limiting accessibility and early motor change quantification. This study assessed the validity of a smartphone app-based FTT, which may expand access and enable more frequent testing.

**Methods:** The cohort was diagnostically diverse, including participants with frontotemporal dementia (FTD), progressive supranuclear palsy (PSP), corticobasal syndrome, primary progressive aphasia, multiple sclerosis, and clinically unimpaired controls. Participants completed a 20-second ALLFTD Mobile App (mApp)-FTT with each hand. Tapping speed metrics were extracted. Participants completed the gold-standard mechanical FTT, a neurologist-administered finger tapping exam, the PSP Rating Scale (PSPRS) and the Unified Parkinson’s Disease Rating Scale (UPDRS). Correlations assessed mApp-FTT and mechanical FTT relationships; regressions evaluated associations with neurologist-rated finger tapping impairment, PSPRS and UPDRS, adjusting for age and sex.

**Results:** The mApp-FTT showed moderate-to-strong correlations with the mechanical FTT (dominant: r=0.63, p<0.001; non-dominant: r=0.55, p<0.001). Taps per second were associated with PSPRS motor severity (dominant hand: std. β=-0.59, 95% CI [-0.91, -0.27], p<0.001) and the UPDRS (dominant hand: std. β=-0.41, 95% CI [-0.82, 0.00], p=0.049). Flight time was modestly associated with neurologist-rated finger tapping impairment (dominant hand: std. β=0.15, 95% CI [0.00, 0.29], p=0.044).

**Conclusion:** These findings support mApp-FTT validity as a measure of motor function across neurodegenerative conditions. Validation in longitudinal and unsupervised remote settings is warranted to understand scalability and evaluate change over time.

## Introduction

Motor impairments, including reduced speed, strength, dexterity, and coordination, are common across neurological disorders [1]. They are especially prominent in neurodegenerative diseases such as Parkinson’s disease (PD) [2] and amyotrophic lateral sclerosis (ALS) [3], demyelinating conditions such as multiple sclerosis (MS) [4], and other neurodegenerative diseases that affect the motor system including frontotemporal dementia (FTD). Motor impairments affect daily functioning and quality of life, and their development often signals disease onset or progression [1]. Accurate quantification of motor speed is therefore an important component of neurological and neuropsychological evaluations.

The finger tapping test (FTT), also referred to as the finger oscillation test, is a mechanical measure of motor speed originally included as part of the Halstead–Reitan Neuropsychological Test Battery in 1947. It has been used for decades in clinical and research settings as a quantitative motor assessment tool, and survey data have placed it in the top 10 most commonly used tests in neuropsychological assessment [5, 6]. This task has demonstrated strong test-retest reliability and validity across multiple clinical populations, including PD [7], MS [8–10], AD [11, 12], and individuals with frontal lobe damage [13, 14]. The finger tapping test is also sensitive to lateralized motor deficits [11, 15, 16].

The traditional FTT task and evaluations by a neurologist require in-person clinical visits. These clinical examinations and rating scales are costly, time-limited, and typically administered infrequently (e.g., annually) [7, 17]. This creates barriers for patients in underserved or rural settings, and limits the ability to capture the precise temporal evolution of motor change, which may progress over relatively short timescales in progressive supranuclear palsy (PSP) and corticobasal syndrome (CBS) [18]. Moreover, as the field more regularly incorporates remote data collection methods and considers decentralized clinical trials, digital tools for quantitative, remote motor assessment will be essential for tracking disease progression and treatment response [19, 20]. More recently, the finger tapping task has been digitized for use on smartphones and tablets, which can enable remote testing and automated recording of tapping behavior [16, 21–25]. Digitized finger tapping tasks in PD discriminate between on/off levodopa status and are associated with disease severity [22, 25–27]. In patients with MS, they are associated with upper-limb dysfunction and Expanded Disability Status Scale scores [8–10]. Less is known about how these tasks perform in FTD syndromes, which include the behavioral variant of FTD (bvFTD), language syndromes known collectively as primary progressive aphasia (PPA) [28], and three motor-predominant syndromes: PSP, CBS and ALS [29]. These three motor syndromes can affect dexterity, speed, and strength of the upper extremities. As FTD progresses, even those with primary behavioral and language syndromes also commonly develop a range of motor features, including parkinsonism, weakness, and dystonia [30]. Motor features are common in both sporadic and familial forms of FTD [30, 31].

To explore digitized quantification of finger tapping in FTD and other disorders, researchers from the ARTFL-LEFFTDS Longitudinal FTLD (ALLFTD) consortium partnered with Datacubed Health to develop the ALLFTD-mobile App (ALLFTD-mAPP), a smartphone application designed to assess cognitive, speech, and motor changes in FTLD and other neurological diseases. Initial studies of this technology focused on the cognitive tasks, demonstrating feasibility and acceptability [32, 33], as well as reliability and validity [34, 35]. The ALLFTD-mApp also includes tasks of gait, balance, and a digitized FTT. Most of our knowledge of motor features of FTD is derived from qualitative clinician observation and neurological examination, whereas digital quantitative assessment tools have been relatively less developed.

In this study, we sought to validate the ALLFTD-mApp-FTT. Although this task is available on smartphones and tablets and designed for remote deployment, to initially investigate task validity under optimal conditions, this study reports on data gathered during an in-person visit using a study-provided smartphone under the observation of trained clinical research coordinators. Participants with a variety of clinical syndromes, predominantly in the FTD spectrum, completed neurological examinations, clinical scales, smartphone FTT, and a mechanical FTT. To investigate construct validity of the app-based version of the FTT, we compared the mApp-FTT to these measures. We evaluated multiple mApp-FTT outcomes including total taps, flight time, hold time, and taps per second in a neurologically diverse cohort, including participants with a range of FTD diagnoses.

## Methods

### Participants

Participants (n=69) were enrolled at the UCSF Weill Institute for Neurosciences for an in-person digital phenotyping study; one portion of this study involved completing tasks under research coordinator supervision, including a mechanical FTT and smartphone-based cognitive and motor tests. Participants were referred from several longitudinal studies, including the UCSF Alzheimer’s Disease Research Center (ADRC), the Frontotemporal Dementia Program Project Grant (PPG; P01 AG019724), the ALLFTD study (U19 AG063911; NCT04363684), the Brain Aging Network for Cognitive Health (BrANCH) study (R01 AG032289), and the UCSF BrainWalk study. Recruitment occurred between January 2023 and July 2025.

Inclusion criteria were: (1) age ≥ 18 years; (2) sufficient fluency in English; (3) completion of a neurological assessment; (4) participating in other UCSF research studies; and (5) presence of a diagnosed neurodegenerative disorder or clinically unimpaired. Diagnoses were made as part of a multidisciplinary case conference using established diagnostic criteria [36–38].

### ALLFTD Mobile App FTT

ALLFTD investigators partnered with Datacubed Health (www.datacubed.com) to develop the ALLFTD-mApp. The app is compatible with iOS (version 11+) and Android (version 6+) devices. All collected data are securely maintained on Amazon Web Service servers that meet HIPAA, GDPR, and FDA 21 CFR Part 11 compliance requirements [32]. The app includes surveys and measures of cognition and motor function. It captures audio samples while participants respond to speech and language prompts [34], and the app passively collects data through phone sensors, such as step count and battery usage [33]. The focus of this study was the mApp-FTT (Figure 1). Participants completed the task on a study-provided iPhone (SE, version 11).

**Figure 1.**
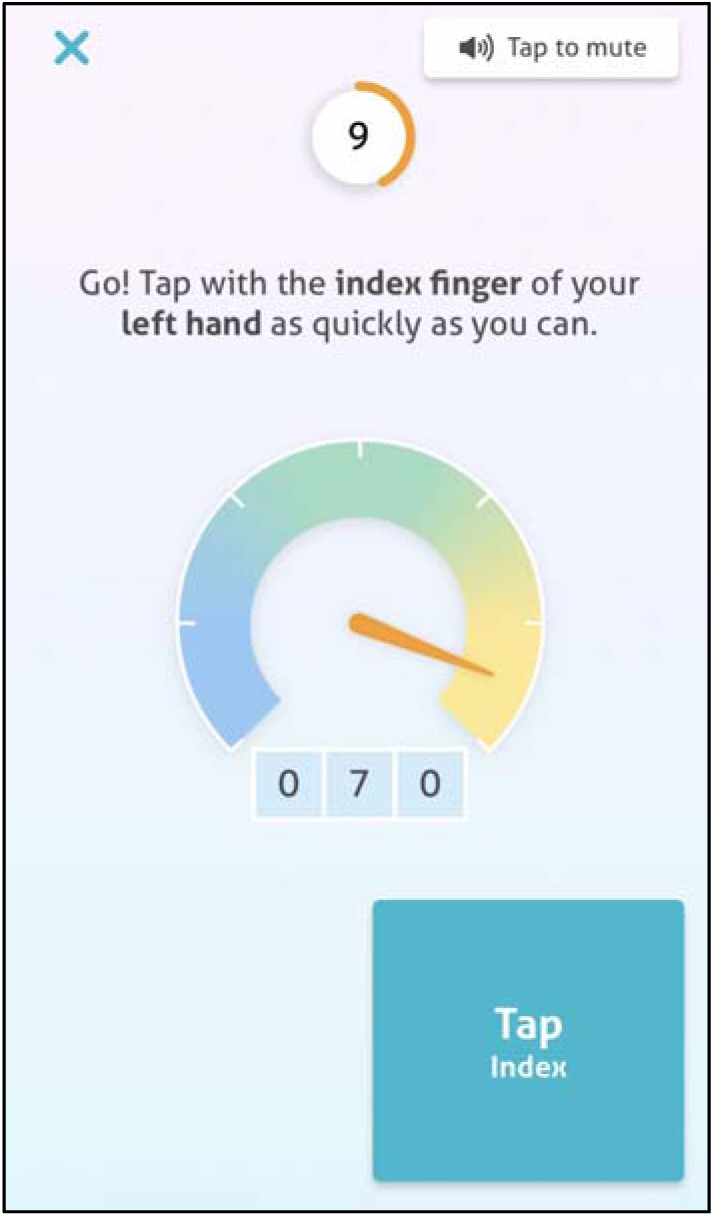
Screenshot of the ALLFTD Mobile App finger-tapping test. *Participants are instructed to tap as quickly as possible using their index finger for a 20-second trial. The interface displays a “speedometer” which indicates tapping speed. A running count of taps is also displayed. Tapping is completed separately for each hand.*

Participants identified their dominant hand on the app and were asked to complete the task with each hand separately. They were given the option to skip a hand if they were unable to complete the task with that hand. Next, they were instructed to tap as quickly as possible for 20 seconds with their index finger, while keeping their palm on the table. In an early version of the task, trials lasted 30 seconds; for those participants (n=2), their data were truncated at 20 seconds to ensure consistency across the sample. Metrics derived from the mApp-FTT also included flight time and hold time. Flight time was defined as the inter-tap interval, measured from the release of one tap to the initiation of the next. Hold time was defined as the duration from initial screen contact to tap release. Implausibly short flight and hold time values suggestive of measurement error were excluded prior to metric calculation, consistent with prior smartphone-based tapping studies [26, 39, 40]. Thresholds were defined a priori (flight time <0.05 seconds; hold time <0.01 seconds). Similarly, a threshold of ±3 standard deviations from each participant’s trial-level median was implemented to remove trials of excessively long duration that may be suggestive of poor task engagement or errors. The remaining valid flight times and hold times were added together to calculate the total tapping time for that trial. Taps per second were calculated as the total remaining number of taps divided by this adjusted trial duration. Participants with less than 85% of taps remaining were excluded from this study (n=2). Smartphone mApp-FTT outcomes were taps per second, total tap count, average flight time, and average hold time.

### Clinical Assessments

#### Mechanical FTT

Participants completed the mechanical Halstead–Reitan FTT at the same appointment as their mApp-FTT. A modified version of a brief administration paradigm was used [7], consisting of one practice trial followed by five 10-second trials for each hand, during which index-finger taps were recorded by a study coordinator using a mechanical counter. Participants completed all trials with the dominant hand first, followed by all trials with the non-dominant hand. The outcome measure was the mean number of taps across the five trials. A separate score was calculated for each hand.

#### Neurological Exam Finger Tapping (MDS-UPDRS)

Finger tapping was assessed by a neurologist using the Movement Disorder Society-Unified Parkinson’s Disease Rating Scale (MDS-UPDRS) Part III, finger tapping item. Neurologists performing the UPDRS exam were blinded to smartphone tapping results. Participants tapped their index finger to their thumb as quickly as possible for 10 seconds. Performance was rated on a 0-4 scale, with 0 indicating normal ability and 4 indicating severe impairment. For analyses, scores were examined as both continuous and dichotomized variables with dichotomized scores defined as unimpaired (score = 0) or impaired (score > 0). Dominant and non-dominant hands were assessed separately.

#### Unified Parkinson’s Disease Rating Scale (UPDRS) Total Score

Participants also completed the Movement Disorder Society-Unified Parkinson’s Disease Rating Scale (UPDRS), which assesses motor and non-motor symptoms across multiple domains. The score used in analyses reflects overall symptom severity, with higher scores indicating greater impairment. Scores ranged from 0 to 32 in this sample.

#### Progressive Supranuclear Palsy Rating Scale (PSPRS)

A neurologist administered the PSPRS, which quantifies the severity of motor impairment across multiple domains [41]. Total scores range from 0 to 100, with higher scores indicating greater motor and clinical impairment.

### Statistical Analysis

Participant characteristics were summarized using descriptive statistics. To assess construct validity, Pearson correlation coefficients were calculated to assess the strength of association between smartphone-based finger tapping metrics and mechanical FTT performance. Associations between smartphone-based metrics and clinical measures were examined using linear regression models, with smartphone metrics as independent variables and clinical measures as dependent variables, adjusting for age and sex. Separate regressions were conducted for dominant and non-dominant hands and for each clinical outcome (mechanical FTT, neurologist-rated finger tapping, PSPRS total score, and UPDRS total score). Each smartphone metric (e.g., taps per second, flight time) was entered in a separate model, with taps per second considered as the primary variable, and other analyses considered exploratory. Standardized regression coefficients (std β) were estimated by z-standardizing continuous predictor and outcome variables (mean = 0, SD = 1) prior to model fitting, allowing comparison of effect sizes across tapping metrics.

ANCOVAs were used to investigate smartphone-based metric performance across diagnostic groups, covarying for age and sex. A priori pairwise comparisons focused on clinically unimpaired participants versus (1) a FTLD motor condition group (PSP and CBS) and (2) a MS group. Unadjusted contrasts were evaluated first, followed by Tukey HSD correction to account for multiple comparisons (2 planned comparisons per metric).

Analyses were conducted using R (v4.3.2; RStudio 2025.05.0) and Python (Anaconda, Jupyter Notebook).

## Results

### Participant Characteristics

A total of 69 participants were included in the study (Table 1). The mean age was 66.6 years (SD = 11.9), and the median education was 18 years (IQR = 16-18). More than half of the sample were male (n = 39, 56.5%), and the majority (n = 61, 88.4%) were right-hand dominant. Most participants identified as White (n = 50, 72.5%).

**Table 1.** Participant Characteristics. *Values are presented as mean (SD) or median (IQR) for continuous variables and frequency (%) for categorical variables. Abbreviations: SD = standard deviation; IQR = interquartile range; bvFTD = behavioral variant frontotemporal dementia (FTD); nfvPPA = nonfluent/agrammatic variant primary progressive aphasia (PPA); MS = multiple sclerosis; MCBI/MCI = mild cognitive/behavioral impairment; PSP = progressive supranuclear palsy; CBS = corticobasal syndrome; svPPA = semantic variant PPA; sbvFTD = semantic behavioral variant FTD; lvPPA = logopenic variant PPA; TBI = traumatic brain injury. We did not describe breakdown of race/ethnic categories further due to the small number of participants that could be identifying. Disease severity was characterized using the Progressive Supranuclear Palsy Rating Scale (PSPRS) and the Clinical Dementia Rating® plus National Alzheimer’s Coordinating Center Frontotemporal Lobar Degeneration module (CDR® plus NACC FTLD).*

| Characteristic |  |
| --- | --- |
| **Total Participants, n** | **69** |
| **Age (mean ± SD)** | 66.6 ± 11.9 |
| **Education in Years (median [IQR])** | 18.0 [16.0–18.0] |
| **Male Sex, n (%)** | 39 (56.5) |
| **Right Handedness, n (%)** | 61 (88.4) |
| **White, n (%)** | 50 (72.5) |
| **Clinical Phenotype, n (%)** |  |
| bvFTD | 16 (23.2) |
| nfvPPA | 10 (14.5) |
| Clinically unimpaired | 11 (15.9) |
| PSP | 6 (8.7) |
| MCBI/MCI | 3 (4.3) |
| MS | 7 (10.1) |
| CBS | 7 (10.1) |
| svPPA | 2 (2.9) |
| sbvFTD | 2 (2.9) |
| lvPPA | 4 (5.8) |
| TBI | 1 (1.4) |
| **PSPRS total (mean ± SD)** | 6.9 ± 8.2 |
| **PSPRS total > 0, n (%)** | 33 (47.8) |
| **CDR® plus NACC FTLD Global Score, n (%)** |  |
| 0 | 9 (13.0) |
| 0.5 | 16 (23.2) |
| 1+ | 31 (44.9) |
| Missing | 13 (18.8) |

The diagnostic distribution was diverse, encompassing both FTD syndromes and other neurological conditions. The most common clinical phenotypes were bvFTD (n = 16, 23.2%), nonfluent variant primary progressive aphasia (nfvPPA; n = 10, 14.5%), and clinically unimpaired individuals (n = 11, 15.9%). Other subgroups included PSP (n = 6, 8.7%), mild cognitive/behavioral impairment (MCBI; n = 3, 4.3%), MS (n = 7, 10.1%), CBS (n = 7, 10.1%), semantic variant PPA (svPPA; n = 2, 2.9%), semantic-behavioral variant FTD (sbvFTD; n = 2, 2.9%), and logopenic variant PPA (lvPPA; n = 4, 5.8%). A single participant had a primary clinical diagnosis of traumatic brain injury (TBI; n = 1, 1.4%).

### Associations with Mechanical Tapping

The mApp-FTT scores correlated strongly with gold-standard mechanical FTT performance: dominant hand taps per second (Figure 2A; r = 0.63, 95% CI [0.43, 0.77], p < 0.001) and total taps (Figure 2C; r = 0.65, 95% CI [0.46, 0.78], p < 0.001); non-dominant hand taps per second (Figure 2B; r = 0.55, 95% CI [0.32, 0.71], p < 0.001) and total taps (Figure 2D; r = 0.57, 95% CI [0.35, 0.73], p < 0.001).

**Figure 2.**
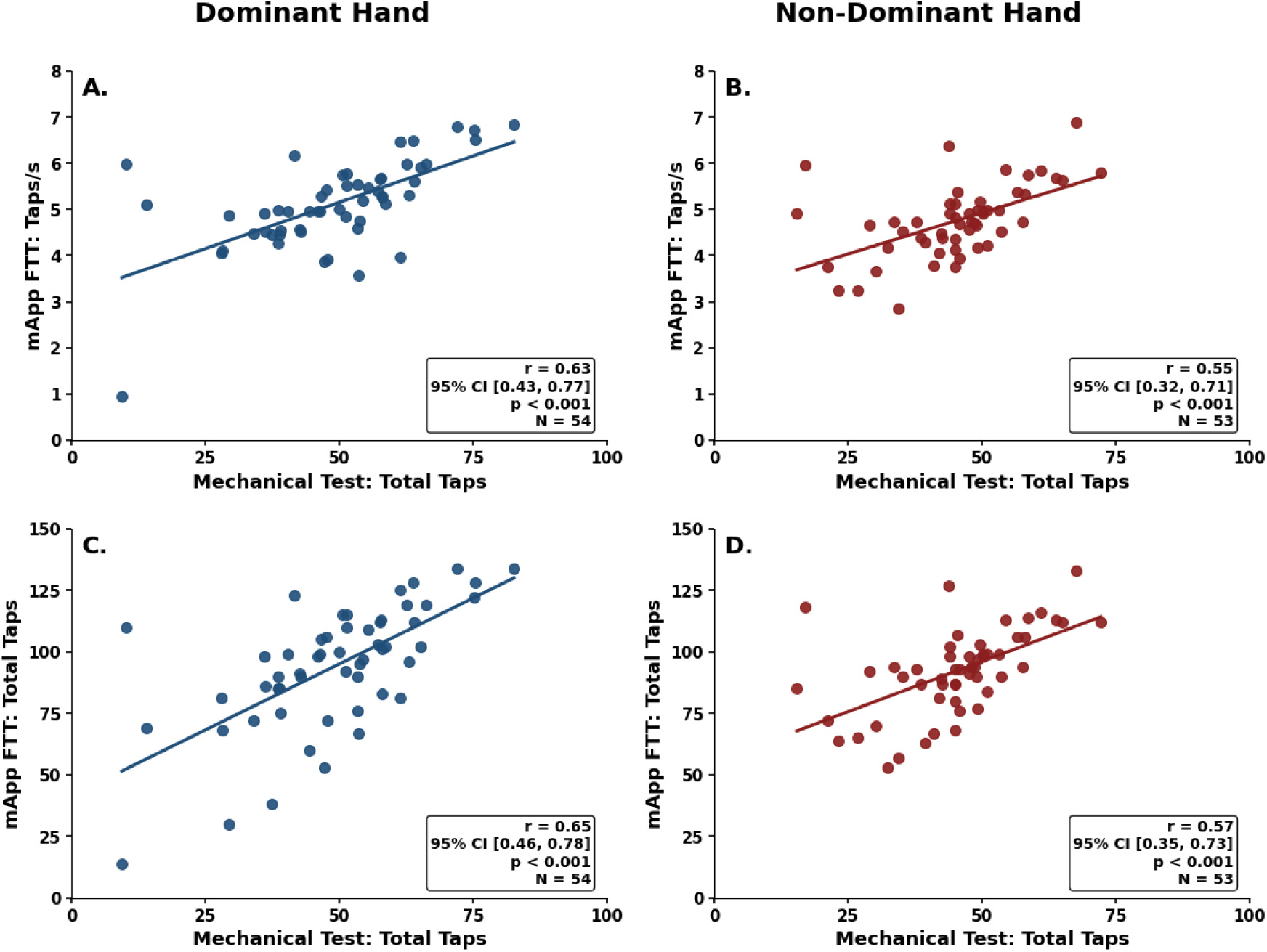
Associations between mechanical and smartphone-based finger tapping. *Scatterplots display the relationship between performance on the gold-standard mechanical finger tapping test and the smartphone-based ALLFTD-mApp finger tapping task. Panels (A, B) display associations between mechanical total taps and smartphone tapping speed (taps/s), and panels (C, D) display associations between mechanical total taps and smartphone total taps. Results are stratified by hand dominance (A, C: dominant hand; B, D: non-dominant hand). Smartphone-derived metrics were calculated following exclusion of implausible inter-tap intervals and contact durations and application of predefined quality control procedures (see Methods). Pearson correlation coefficients are shown with corresponding 95% confidence intervals.*

### Associations With PSPRS and UPDRS

Smartphone tapping metrics were also associated with motor disease severity as measured by the PSPRS (Table 2, Figure 3). For the dominant hand, fewer taps per second were significantly associated with higher PSPRS scores (std. β = -0.59, 95% CI [-0.91, -0.27], p < 0.001), as were fewer total taps (std. β = -0.58, 95% CI [-0.92, -0.24], p = 0.002), longer mean flight times (std. β = 0.68, 95% CI [0.40, 0.95], p < 0.001), and longer mean hold times (std. β = 0.64, 95% CI [0.37, 0.91], p < 0.001). In contrast, non-dominant-hand associations were weaker and not statistically significant (taps per second: std. β = -0.09, 95% CI [-0.48, 0.30], p = 0.655; total taps: std. β = -0.18, 95% CI [-0.56, 0.20], p = 0.346; flight time: std. β = -0.28, 95% CI [-0.76, 0.19], p = 0.235; hold time: std. β = 0.12, 95% CI [-0.24, 0.47], p = 0.499), though the relationships were in the expected direction with the exception of mean flight time. Mechanical finger tapping demonstrated significant associations with PSPRS total score, with effect sizes comparable to those observed for smartphone-based metrics (dominant: std. β = -0.56, 95% CI [-0.86, -0.26], p < 0.001; non-dominant: std. β = -0.33, 95% CI [-0.65, -0.01], p = 0.043).

**Table 2.**
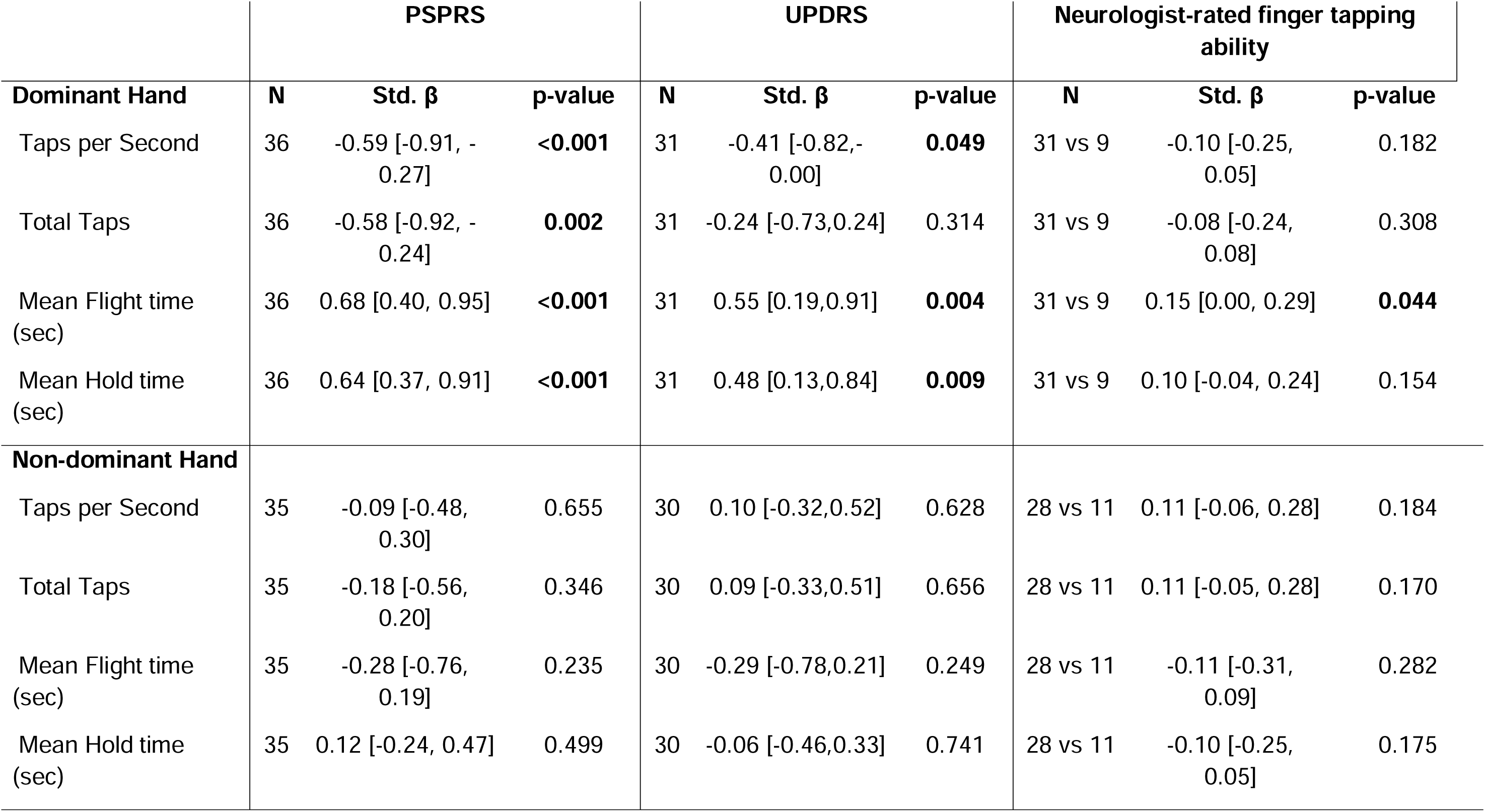
Associations between smartphone tapping metrics and clinical measures of motor impairment. *Participants were categorized as impaired or not impaired based on neurologist rating of performance on an independent bedside finger-tapping test and were also evaluated using the Progressive Supranuclear Palsy Rating Scale (PSPRS) and the Unified Parkinson’s Disease Rating Scale (UPDRS). Smartphone tapping metrics were compared with these clinical measures, stratified by hand dominance. Standardized beta coefficients (adjusted for age and sex) are reported with 95% confidence intervals [CI]. Bold p-values with stars indicate statistical significance. Note: Differences in sample size across smartphone metrics (N = 35-40) reflect varying completeness of timestamp data required to compute rate- and time-based variables. Total tap counts were available for slightly more participants than duration-dependent measures (taps per second, flight, and hold times).*

**Figure 3.**
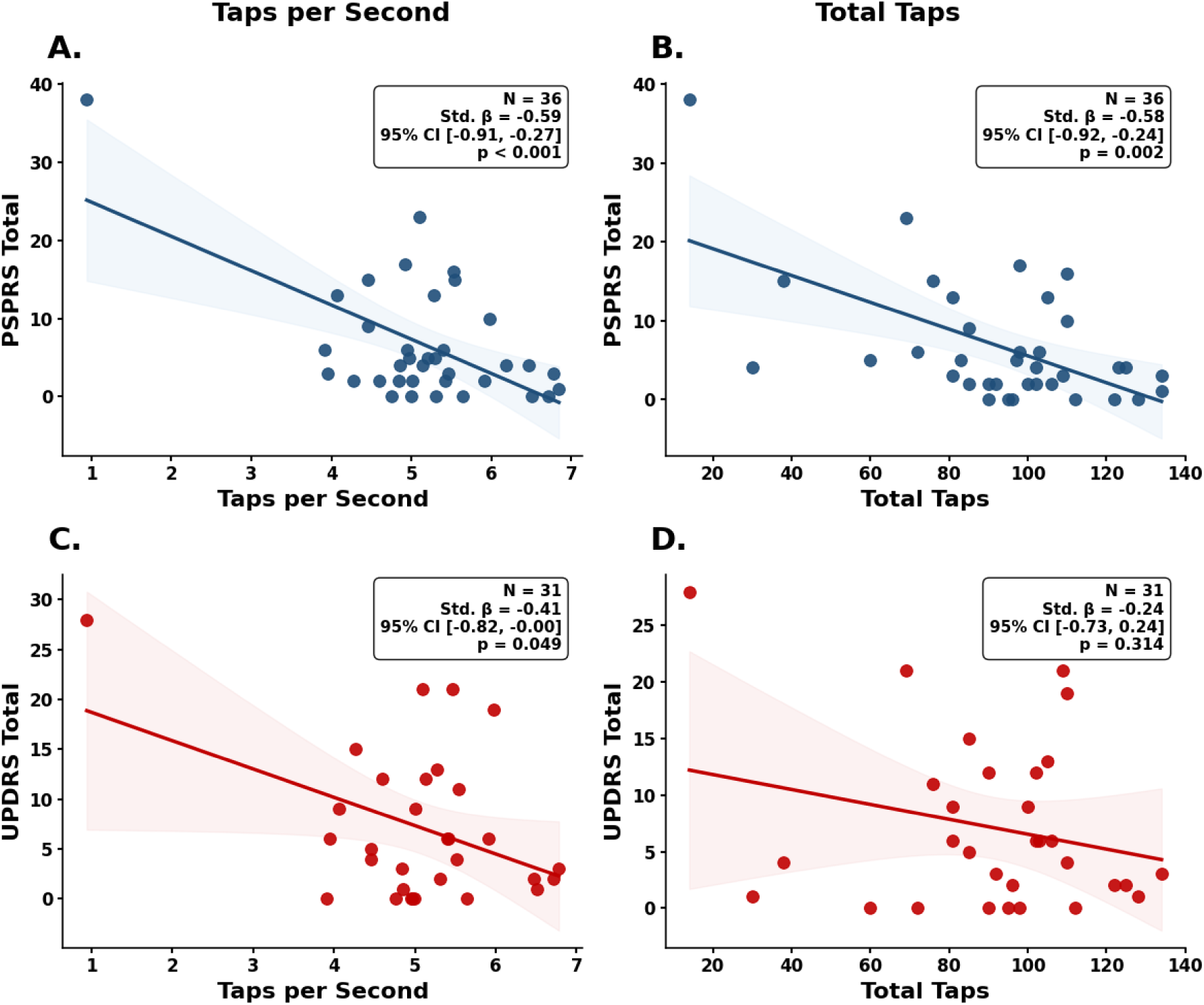
Association between smartphone-based finger tapping performance and PSPRS and UPDRS total scores (dominant hand). *Scatterplots show the relationship between smartphone-derived tapping metrics and (A, B) Progressive Supranuclear Palsy Rating Scale (PSPRS) total scores and (C, D) Unified Parkinson’s Disease Rating Scale (UPDRS) total scores. Panels display taps per second (A, C) and total taps (B, D). All models are adjusted for age and sex and are presented with standardized β coefficients. Blue regression lines represent PSPRS models and red regression lines represent UPDRS models, with shaded areas indicating 95% confidence intervals.*

A similar pattern of associations was observed when smartphone tapping metrics were compared with UPDRS total scores (Table 2, Figure 3). For the dominant hand, fewer taps per second were significantly associated with higher UPDRS scores (std. β = -0.41, 95% CI [-0.82, 0.00], p = 0.049), as were longer mean flight times (std. β = 0.55, 95% CI [0.19, 0.91], p = 0.004) and longer mean hold times (std. β = 0.48, 95% CI [0.13, 0.84], p = 0.009), whereas total taps were not (std. β = -0.24, 95% CI [-0.73, 0.24], p = 0.314). In contrast, non-dominant-hand associations were weaker and not statistically significant (taps per second: std. β = 0.10, 95% CI [-0.32, 0.52], p = 0.628; total taps: std. β = 0.09, 95% CI [-0.33, 0.51], p = 0.656; flight time: std. β = -0.29, 95% CI [-0.78, 0.21], p = 0.249; hold time: std. β = - 0.06, 95% CI [-0.46, 0.33], p = 0.741). Mechanical finger tapping demonstrated significant associations with UPDRS total score, with effect sizes comparable to those observed for smartphone-based metrics (dominant: std. β = -0.45, 95% CI [-0.80, -0.11], p = 0.012; non-dominant: std. β = -0.32, 95% CI [-0.67, 0.02], p = 0.065).

### Associations With Neurologist-Rated Finger Tapping

For the dominant hand, longer mean flight times (std. β = 0.15, 95% CI [0.00, 0.29], p = 0.044) were significantly associated with neurologist-rated finger-tapping impairment. Taps per second (std. β = -0.10, 95% CI [-0.25, 0.05], p = 0.182) were not significantly associated with neurologist-rated finger-tapping impairment, nor were fewer total smartphone taps (std. β = -0.08, 95% CI [-0.24, 0.08], p = 0.308), or longer mean hold times (std. β = 0.10, 95% CI [-0.04, 0.24], p = 0.154), though all relationships were in the expected direction. In contrast, associations for non-dominant hand metrics were smaller in magnitude and not statistically significant, with wide confidence intervals that included the null (Table 2).

### Smartphone-based Metrics Across Diagnostic Groups

After adjusting for age and sex, dominant hand taps per second did not differ significantly across groups but trended toward group differences (ANCOVA p = 0.083), whereas total taps differed significantly (ANCOVA p = 0.042; Figure 4). Prespecified comparisons demonstrated reduced taps per second in participants with FTLD motor syndromes (PSP and CBS) compared with clinically unimpaired individuals (mean difference = -1.23, 95% CI [-2.42, -0.04], Tukey-adjusted p = 0.038), with total taps at the threshold of significance in the expected direction (mean difference = -29.40, 95% CI [-58.82, 0.02], Tukey-adjusted p = 0.050). There were no significant differences between participants with MS and clinically unimpaired participants (ps > 0.05). No significant differences across diagnostic groups were found for non-dominant-hand total taps, taps per second, hold time, or flight times (ps > 0.05).

**Figure 4.**
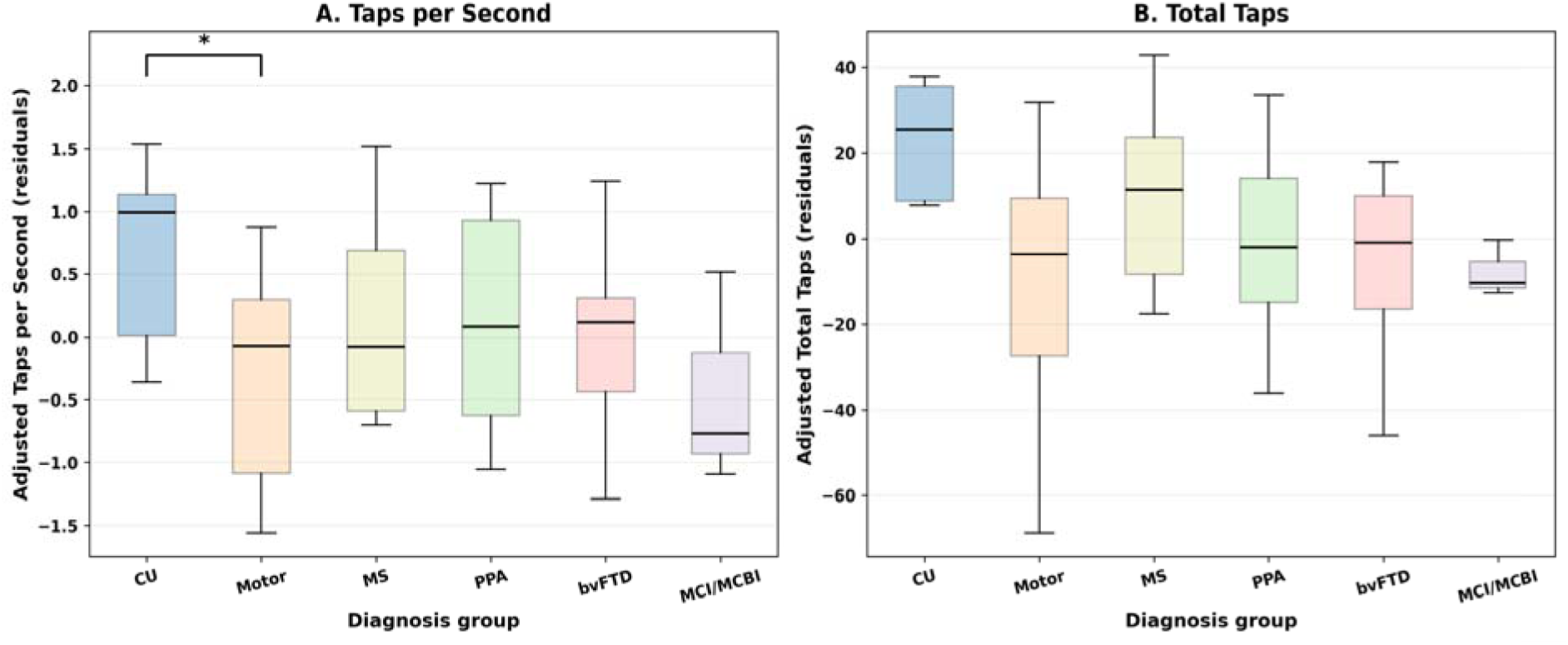
Dominant-hand smartphone finger tapping across diagnostic groups. *(A) Tapping rate (taps per second) and (B) total taps are shown by diagnosis, including clinically unimpaired (CU) participants, motor syndromes (progressive supranuclear palsy [PSP] and corticobasal syndrome [CBS]), multiple sclerosis (MS), primary progressive aphasia (PPA), behavioral variant frontotemporal dementia (bvFTD), and mild cognitive impairment/mild cognitive behavioral impairment (MCI/MCBI). Values were adjusted for age and sex. Overall group differences were not statistically significant for tapping rate (ANCOVA p=0.083) but were significant for total taps (ANCOVA p=0.042). Post hoc comparisons showed reduced tapping rate in motor syndromes compared with clinically unimpaired participants (Tukey-adjusted p=0.038), with total taps at the threshold of significance (Tukey-adjusted p=0.050). *p < 0.05*

## Discussion

This study provides in-clinic validation of a smartphone-based finger-tapping task across a neurologically diverse sample that included those on the FTLD spectrum. Performance on the ALLFTD-mApp-FTT showed strong agreement with gold-standard measures, including the mechanical FTT, PSPRS, and UPDRS. Participants with motor syndromes of PSP and CBS demonstrated slower tapping rates and longer flight and hold times compared to controls, suggesting the mApp-FTT may be useful for detecting motor dysfunction in FTLD. These data were collected during supervised in-clinic completion of this task and support data collection in remote and unsupervised settings.

Mechanical finger tapping is a well-established measure of motor speed and dexterity [7, 42] that is used clinically for many conditions. For example, studies of PD have found moderate-to-strong associations of mechanical FTT with magnitude of motor impairment and reduced tapping performance as compared to controls [40, 44–47]. Here we demonstrate that several metrics from the mApp-FTT are associated with performance on the mechanical FTT, supporting its validity. The strength of the associations was comparable or stronger than those reported in other studies [24, 44], and provides evidence of convergent validity.

The present study also demonstrated the validity of digital finger-tapping tasks in FTLD syndromes through associations of dominant hand performance with clinical ratings of motor symptoms, including the PSPRS, UPDRS, and neurologist bedside rating of finger tapping performance that was collected independently from the smartphone FTT task. The strength of the associations of the mApp-FTT with the PSPRS and the UPDRS were very similar to associations of the mechanical FTT with the PSPRS and the UPDRS, further supporting mApp-FTT clinical utility. These findings build upon similar findings observed in related conditions. In PD, for example, smartphone- and tablet-based FTTs have moderate-to-strong correlations with the UPDRS [24, 26, 47, 48]. Smartphone-based FTTs have also demonstrated discriminative validity in PD, distinguishing patients from healthy controls and differentiating motor states such as on/off phases of levodopa [22, 25, 26]. In longitudinal analyses of PD participants, smartphone FTTs exhibited excellent test-retest reliability [49–53]. In MS, smartphone FTTs correlate with the magnitude of disability as indexed by the Expanded Disability Status Scale [8].

In addition to associations with clinical scales, we also observed differences in tapping performance across diagnostic groups. Our prespecified analyses revealed significant differences between motor-predominant FTLD participants (CBS and PSP) and those who were clinically unimpaired. We also observed slow tapping in some patients with non-motor syndromes, such as bvFTD. Upon further inspection, we discovered that clinicians also scored their bedside finger tapping as impaired, suggesting that the mApp-FTT may be detecting the onset of motor changes even if behavioral features are the earliest and most pronounced features in these participants. Alternately, slower tapping in these participants could reflect the contribution of other cognitive and neuropsychiatric symptoms to tapping performance, including apathy, slowed processing speed, or difficulty sustaining attention. Future work in larger samples will aim to identify patterns in the finger tapping data that may indicate reduced task engagement or cognitive/behavioral impacts on testing.

Despite robust dominant-hand findings, associations between non-dominant hand mApp-FTT metrics and both mechanical finger tapping performance and other clinical measures were weaker than expected and did not reach statistical significance. This pattern of weaker non-dominant-hand associations has also been observed in prior studies involving both mechanical and smartphone-based FTTs [22, 24, 54]. We hypothesize that these differences reflect a combination of dominant-hand motor reserve and familiarity, increased non-dominant-hand variability, and syndrome-specific patterns of upper-limb motor involvement.

There are several strengths to the design of this study. First, conducting the validation in a supervised, in-clinic setting allowed direct comparison of smartphone tapping with mechanical FTT and clinical benchmarks (blinded neurologist ratings, PSPRS, and UPDRS), strengthening interpretability. These findings set the stage for studies of unsupervised data collection in remote environments, which are underway. Second, the cohort was diagnostically diverse, spanning multiple FTLD syndromes as well as non-FTLD conditions, demonstrating feasibility and validity across heterogeneous populations. Third, the study advanced the field by examining novel features that can only be collected using digitized versions of this task; flight and hold times were associated with clinical impairment and both PSPRS and UPDRS severity, consistent with previous PD studies that identified smartphone-derived tapping as sensitive indicators of motor change [22, 25, 54–56].

This work builds upon validation evidence for use of the ALLFTD-mApp in neurodegenerative diseases. The ALLFTD-mApp, built on the Datacubed Health application, also includes tasks of cognition, speech, walking, and balance, making it a comprehensive tool for assessing common neurological symptoms. Prior research has demonstrated feasibility and validated cognitive modules for reaction time and processing speed [35–37]. These findings were replicated in 1,813 adults aged 18-90 years, confirming strong test-retest reliability, cross-platform consistency, and predictable age-related trajectories [57]. In addition, clinical utility was recently demonstrated by showing that performance on these smartphone cognitive tests predicted post-operative delirium [35], and the clinical relevance of passive data collection such as step count and device-use metrics has been established [33]. The app can also be used to record speech samples, which may be sensitive to a range of neurological conditions. This study adds to prior work in smartphone cognitive testing by providing initial validation evidence for digital motor testing in FTD. Together, these results highlight the capacity of the ALLFTD-mApp for integrated digital assessment across cognitive and motor domains.

Several limitations should be acknowledged. The study was conducted in-person, supervised, and exclusively on iOS devices, which may restrict generalizability to fully remote or Android-based implementations. Additional work to validate performance on an Android-based version of this application is underway. The cross-sectional design and modest subgroup sizes limited power for syndrome-specific and longitudinal analyses. Future research should incorporate remote testing, Android devices, and repeated measures to establish test-retest reliability, scalability, and sensitivity to progression.

In conclusion, this study provides evidence supporting the validation of smartphone-based finger tapping to assess upper-limb motor function in a sample of diverse neurological conditions. The ALLFTD-mApp-FTT showed strong associations with mechanical and clinician-rated measures, providing initial evidence of convergent validity. Within the broader suite of ALLFTD-mApp assessments, finger tapping represents a critical motor measure that, combined with other digital tools, may improve diagnosis and monitoring in FTLD and related neurological disorders.

## Data Availability

Data produced in the present study are available upon reasonable request to the authors.

## Contributions

MO, MSC, and AMS contributed to initial manuscript writing. MO, MSC, RF, SD, AS, and AMS contributed to material preparation, data collection and/or analysis. All authors have read and approved the final manuscript. AMS and ALB provided additional funding support and/or helped to develop the smartphone application.

## Ethics approval

All procedures were approved by the UCSF Institutional Review Board (IRB) in accordance with the Declaration of Helsinki.

## Consent to participate / publish

Written informed consent was obtained from all participants, study partners, or legally authorized representatives, as appropriate.

## Data Availability

The datasets generated and/or analyzed during the current study are not publicly available due to privacy reasons but are available from the corresponding author on reasonable request.

## References

1. Erkkinen MG, Kim M-O, Geschwind MD (2018) Clinical Neurology and Epidemiology of the Major Neurodegenerative Diseases. Cold Spring Harb Perspect Biol 10:a033118. 10.1101/cshperspect.a033118

2. Kalia LV, Lang AE (2015) Parkinson’s disease. The Lancet 386:896–912. 10.1016/S0140-6736(14)61393-3

3. Brown RH, Al-Chalabi A (2017) Amyotrophic Lateral Sclerosis. N Engl J Med 377:162–172. 10.1056/NEJMra1603471

4. Reich DS, Lucchinetti CF, Calabresi PA (2018) Multiple Sclerosis. N Engl J Med 378:169–180. 10.1056/NEJMra1401483

5. Rabin LA, Paolillo E, Barr WB (2016) Stability in Test-Usage Practices of Clinical Neuropsychologists in the United States and Canada Over a 10-Year Period: A Follow-Up Survey of INS and NAN Members. Arch Clin Neuropsychol 31:206–230.:

6. Wayne C, Nathan J, Puente A (2000) Psychological test usage: Implications in professional psychology. Prof Psychol Res Pract 31:141–154. 10.1037/0735-7028.31.2.141

7. Castro K (2024) Finger Tapping Test (FTT). In: Clinical Integration of Neuropsychological Test Results. Golden & Bennett, pp 141–146

8. Navarro-López V, Cano-de-la-Cuerda R, Fernández-González P, et al (2024) Reliability and Construct Validity of a Mobile Application for the Finger Tapping Test Evaluation in People with Multiple Sclerosis. BRAIN Sci 14:. 10.3390/brainsci14040407

9. Metzger R, Garrett K, Christensen A, Foley J (2022) Digital Performance Measures Show Sensitivity to Demographic and Disease Characteristicsin a Multiple Sclerosis Cohort Utilizing the MS Care Connect Mobile App in a Real-world Setting. Neurology 98:

10. Beshears K, Koop M, Owen K, et al (2024) Usability and Validation of Smartphone-based Neuroperformance Tests in Multiple Sclerosis. Neurology 102:. 10.1212/WNL.0000000000205817

11. Roalf DR, Rupert P, Mechanic-Hamilton D, et al (2018) Quantitative assessment of finger tapping characteristics in mild cognitive impairment, Alzheimer’s disease, and Parkinson’s disease. J Neurol 265:1365–1375. 10.1007/s00415-018-8841-8

12. Alty J, Goldberg LR, Roccati E, et al (2024) Development of a smartphone screening test for preclinical Alzheimer’s disease and validation across the dementia continuum. BMC Neurol 24:. 10.1186/s12883-024-03609-z

13. Gopal A, Hsu W-Y, Allen DD, Bove R (2022) Remote Assessments of Hand Function in Neurological Disorders: Systematic Review. JMIR Rehabil Assist Technol 9:e33157. 10.2196/33157

14. Dhand A, Mangipudi R, Varshney AS, et al (2025) Assessment of the Sensitivity of a Smartphone App to Assist Patients in the Identification of Stroke and Myocardial Infarction: Cross-Sectional Study. JMIR Form Res 9:e60465. 10.2196/60465

15. Jeppesen Kragh F, Bruun M, Budtz-Jørgensen E, et al (2018) Quantitative Measurements of Motor Function in Alzheimer’s Disease, Frontotemporal Dementia, and Dementia with Lewy Bodies: A Proof-of-Concept Study. Dement Geriatr Cogn Disord 46:168–179. 10.1159/000492860

16. Suzumura S, Osawa A, Kanada Y, et al (2022) Finger Tapping Test for Assessing the Risk of Mild Cognitive Impairment. Hong Kong J Occup Ther 35:137–145. 10.1177/15691861221109872

17. Coravos A, Khozin S, Mandl KD (2019) Developing and adopting safe and effective digital biomarkers to improve patient outcomes. NPJ Digit Med 2:14. 10.1038/s41746-019-0090-4

18. Pressman PS, Miller BL (2014) Diagnosis and Management of Behavioral Variant Frontotemporal Dementia. Biol Psychiatry 75:574–581. 10.1016/j.biopsych.2013.11.006

19. Nevler N, Niehoff D, Gleixner AM, et al (2025) Developing digital health technologies for frontotemporal degeneration. Alzheimers Dement 21:e70082. 10.1002/alz.70082

20. Staffaroni AM, Tsoy E, Taylor J, et al (2020) Digital Cognitive Assessments for Dementia. Pr Neurol 24–25:

21. Surangsrirat D, Sri-iesaranusorn P, Chaiyaroj A, et al (2022) Parkinson’s disease severity clustering based on tapping activity on mobile device. Sci Rep 12:. 10.1038/s41598-022-06572-2

22. Broeder S, Roussos G, De Vleeschhauwer J, et al (2023) A smartphone-based tapping task as a marker of medication response in Parkinson’s disease: a proof of concept study. J Neural Transm 130:937–947. 10.1007/s00702-023-02659-w

23. Lim W-S, Fan S-P, Chiu S-I, et al (2025) Smartphone-derived multidomain features including voice, finger-tapping movement and gait aid early identification of Parkinson’s disease. Npj Park Dis 11:. 10.1038/s41531-025-00953-w

24. Lee CY, Kang SJ, Hong S-K, et al (2016) A Validation Study of a Smartphone-Based Finger Tapping Application for Quantitative Assessment of Bradykinesia in Parkinson’s Disease. PLOS ONE 11:e0158852. 10.1371/journal.pone.0158852

25. Thijssen E, Makai[Bölöni S, Van Brummelen E, et al (2022) A Placebo[Controlled Study to Assess the Sensitivity of Finger Tapping to Medication Effects in Parkinson’s Disease. Mov Disord Clin Pract 9:1074–1084. 10.1002/mdc3.13563

26. Zhan A, Mohan S, Tarolli C, et al (2018) Using Smartphones and Machine Learning to Quantify Parkinson Disease Severity: The Mobile Parkinson Disease Score. JAMA Neurol 75:876. 10.1001/jamaneurol.2018.0809

27. Fay-Karmon T, Galor N, Heimler B, et al (2024) Home-based monitoring of persons with advanced Parkinson’s disease using smartwatch-smartphone technology. Sci Rep 14:9. 10.1038/s41598-023-48209-y

28. Rascovsky K, Hodges JR, Knopman D, et al (2011) Sensitivity of revised diagnostic criteria for the behavioural variant of frontotemporal dementia. Brain 134:2456–2477. 10.1093/brain/awr179

29. Antonioni A, Raho EM, Lopriore P, et al (2023) Frontotemporal Dementia, Where Do We Stand? A Narrative Review. Int J Mol Sci 24:11732. 10.3390/ijms241411732

30. Onyike CU, Diehl-Schmid J (2013) The epidemiology of frontotemporal dementia. Int Rev Psychiatry 25:130–137. 10.3109/09540261.2013.776523

31. Tipton PW, Deutschlaender AB, Savica R, et al (2022) Differences in Motor Features of *C9orf72*, *MAPT*, or *GRN* Variant Carriers With Familial Frontotemporal Lobar Degeneration. Neurology 99:. 10.1212/WNL.0000000000200860

32. Taylor JC, Heuer HW, Clark AL, et al (2023) Feasibility and acceptability of remote smartphone cognitive testing in frontotemporal dementia research. Alzheimers Dement Diagn Assess Dis Monit 15:e12423. 10.1002/dad2.12423

33. Paolillo EW, Casaletto KB, Clark AL, et al (2024) Examining Associations Between Smartphone Use and Clinical Severity in Frontotemporal Dementia: Proof-of-Concept Study. JMIR Aging 7:e52831. 10.2196/52831

34. Staffaroni AM, Clark AL, Taylor JC, et al (2024) Reliability and Validity of Smartphone Cognitive Testing for Frontotemporal Lobar Degeneration. JAMA Netw Open 7:e244266. 10.1001/jamanetworkopen.2024.4266

35. Barreto Chang OL, Wise AB, Sinha A, et al (2025) Remote Cognitive Testing for Detection of Baseline Cognitive Impairment and Prediction of Postoperative Delirium Risk: A Pilot Study. Anesth Analg. 10.1213/ANE.0000000000007843

36. Morris JC (2012) Revised Criteria for Mild Cognitive Impairment May Compromise the Diagnosis of Alzheimer Disease Dementia. Arch Neurol 69:700–708. 10.1001/archneurol.2011.3152

37. Besser L, Kukull W, Knopman DS, et al (2018) Version 3 of the National Alzheimer’s Coordinating Center’s Uniform Data Set. Alzheimer Dis Assoc Disord 32:351. 10.1097/WAD.0000000000000279

38. Albert MS, DeKosky ST, Dickson D, et al (2011) The diagnosis of mild cognitive impairment due to Alzheimer’s disease: Recommendations from the National Institute on Aging-Alzheimer’s Association workgroups on diagnostic guidelines for Alzheimer’s disease. Alzheimers Dement J Alzheimers Assoc 7:270–279. 10.1016/j.jalz.2011.03.008

39. Espay AJ, Giuffrida JP, Chen R, et al (2011) Differential response of speed, amplitude, and rhythm to dopaminergic medications in Parkinson’s disease. Mov Disord Off J Mov Disord Soc 26:2504–2508. 10.1002/mds.23893

40. Arora S, Venkataraman V, Zhan A, et al (2015) Detecting and monitoring the symptoms of Parkinson’s disease using smartphones: A pilot study. Parkinsonism Relat Disord 21:650–653. 10.1016/j.parkreldis.2015.02.026

41. Golbe LI, Ohman-Strickland PA (2007) A clinical rating scale for progressive supranuclear palsy. Brain 130:1552–1565. 10.1093/brain/awm032

42. Coleman AR, Moberg PJ, Ragland JD, Gur RC (1997) Comparison of the Halstead-Reitan and Infrared Light Beam Finger Tappers. Assessment 4:277–286. 10.1177/107319119700400307

43. Shimoyama I (1990) The Finger-Tapping Test: A Quantitative Analysis. Arch Neurol 47:681. 10.1001/archneur.1990.00530060095025

44. Christianson MK, Leathem J (2004) Development and Standardisation of the Computerised Finger Tapping Test: Comparison with Other Finger Tapping Instruments. N Z J Psychol 44:

45. Dunnewold RJW, Jacobi CE, Van Hilten JJ (1997) Quantitative assessment of bradykinesia in patients with parkinson’s disease. J Neurosci Methods 74:107–112. 10.1016/S0165-0270(97)02254-1

46. Taylor Tavares AL, Jefferis GSXE, Koop M, et al (2005) Quantitative measurements of alternating finger tapping in Parkinson’s disease correlate with UPDRS motor disability and reveal the improvement in fine motor control from medication and deep brain stimulation. Mov Disord 20:1286–1298. 10.1002/mds.20556

47. Simonet C, Galmes MA, Lambert C, et al (2021) Slow Motion Analysis of Repetitive Tapping (SMART) Test: Measuring Bradykinesia in Recently Diagnosed Parkinson’s Disease and Idiopathic Anosmia. J Park Dis 11:1901–1915. 10.3233/jpd-212683

48. Guo W-H, Yang X-D, Ruan Z, et al (2025) Early detection of Parkinson’s disease: Machine learning-based prediction of UPDRS Part III scores in de novo patients using smartphone assessments. J Park Dis 1877718X251359494. 10.1177/1877718X251359494

49. Adams JL, Waddell EM, Chunga N, Quinn L (2023) Digital Measures in Huntington’s Disease. Contemp Clin Neurosci Part F1569:433–457. 10.1007/978-3-031-32815-2_18

50. Waddell EM, Dinesh K, Spear KL, et al (2021) GEORGE®: A Pilot Study of a Smartphone Application for Huntington’s Disease. J Huntingt Dis 10:293–301. 10.3233/JHD-200452

51. Adams JL, Kangarloo T, Tracey B, et al (2023) Using a smartwatch and smartphone to assess early Parkinson’s disease in the WATCH-PD study. Npj Park Dis 9:. 10.1038/s41531-023-00497-x

52. Kontz M, Wattendorf S, Thieken F, et al (2023) A smartphone-based approach to continuous monitoring of Parkinson’s disease patients. Biomed Tech 68:. 10.1515/bmte-2023-2001

53. Lipsmeier F, Simillion C, Bamdadian A, et al (2022) A Remote Digital Monitoring Platform to Assess Cognitive and Motor Symptoms in Huntington Disease: Cross-sectional Validation Study. J Med Internet Res 24:e32997. 10.2196/32997

54. Adams JL, Kangarloo T, Gong Y, et al (2024) Using a smartwatch and smartphone to assess early Parkinson’s disease in the WATCH-PD study over 12 months. Npj Park Dis 10:. 10.1038/s41531-024-00721-2

55. Makai-Bölöni S, Thijssen E, Van Brummelen EMJ, et al (2021) Touchscreen-based finger tapping: Repeatability and configuration effects on tapping performance. PLoS ONE 16:. 10.1371/journal.pone.0260783

56. Lipsmeier F, Taylor KI, Kilchenmann T, et al (2018) Evaluation of smartphone[based testing to generate exploratory outcome measures in a phase 1 Parkinson’s disease clinical trial. Mov Disord 33:1287–1297. 10.1002/mds.27376

57. Dhanam S, Sanderson-Cimino M, Taylor JC, et al (2025) Remote, self-administered, smartphone cognitive testing in a registry-based cohort: Feasibility, reliability, and validity findings. 2025.10.28.25338686

